# Strategies for communicating health evidence to health policymakers and the population. Scoping review protocol

**DOI:** 10.1101/2021.11.04.21265922

**Authors:** Ana Luiza Cabrera Martimbianco, Rafael Leite Pacheco, Ângela Maria Bagattini, Roberta de Fátima Carreira Moreira, Rachel Riera

**Author notes:** **Correponding author:** Rafael Leite Pacheco, Rua Barata Ribeiro, 142- Bela Vista — São Paulo (SP) — Zip Code 01308-000.

## Abstract

Evidence-based health information is provided by evidence synthesis and health technology assessments. Nevertheless, this information is complex for public understanding, pointing to the need to disseminate clearly. This scoping review aims to identify different strategies for communicating health evidence to policymakers and the general population. A scoping review will be conducted following the Joanna Briggs Institute Manual for Scoping Reviews. This comprehensive mapping will contribute to identifying the literature on health evidence-based information, identify the most appropriate approaches for each audience, and the literature gaps to guide future studies.

## Introduction

Evidence synthesis and health technology assessments provide evidence-based information on health conditions, interventions, and procedures, to meet the needs of health professionals, patients and public or private health policymakers. Nevertheless, evidence obtained from scientific studies, mainly from systematic reviews and other syntheses of multiple studies, is complex and often difficult for public understanding (McCormack, 2013). For this reason, health evidence needs to be communicated and disseminated in such a way that it is clearly understood by decision-makers, especially in contexts that require quick responses.

Health evidence communication strategies aim to increase the understanding of the information transmitted and should encompass products that meet the needs of the health policymakers (given the demands of health care services) and the population (reliable information based on the best available scientific evidence).

Therefore, expanding investment in communication allows identifying the best strategies to overcome the barrier among health care evidence, health policymakers and population. Clear communication and active dissemination of health evidence to all relevant stakeholders in easy-to-understand and accessible formats are essential to raising awareness about the use of scientific evidence, informing and influencing individual and community health-related decisions (McCormack, 2013).

In the last decade, digital and social media advancement has reformulated the concept of communication in health, allowing for new, direct, and influential communication platforms and channels between researchers and the public (Brownell, 2013; Fontaine, 2018). Several strategies, such as plain language summaries and infographics, have been developed and tried out for this purpose. Thus, a comprehensive mapping is needed to identify the nature and extent of the literature related to the fundamental concept of these approaches, identify, and assess the most appropriate approaches for each audience and context, and the gaps in the literature. The purpose of this scoping review is to identify and assess the different strategies for communicating health evidence (content and formats) to health policymakers and the population.

## Methods

We will conduct a scoping review following the Joanna Briggs Institute Manual for Scoping Reviews (Peters, 2020). In addition, the review report will follow the recommendations of Preferred Reporting Items for Systematic Reviews and Meta-Analyses - extension for scoping reviews (PRISMA-ScR) (Tricco, 2018).

### Methods to involve society and stakeholders in the project

Stakeholders were consulted throughout this review protocol development to increase the applicability of the review findings and support the communication and translation of the results to be used by society. We considered consulting the following stakeholders:

- Consumers (health policymakers, healthcare professionals and patients): to contribute to the topic refinement, clarify definitions, present their understanding of the concepts, strategies and programs identified by this evidence synthesis. This process aims to increase the relevance of the scoping review and reduce the potential for research waste.
- Content experts: to assess whether the review is relevant to the field of knowledge and help identify resources that the preliminary search on the data sources may not identify.
- Methodology experts: to support the scoping review development and conduction and to answer any methodological questions that may arise during this process.
- Information specialists support the definition of a relevant search strategy and identify databases relevant to this topic.

The research question of this review was structured using the acronym PCC as follows:

- P (population, condition): not applicable.
- C (concept): strategies for communicating health evidence to health policymakers and the population.
- C (context): health policies.

### Criteria for inclusion of studies according to the components of the PCC acronym

- P (population, condition): not applicable
- C (concept): strategies for communicating health evidence to health policymakers and/or the population. We will consider any strategy or program focusing on the translation, adaptation or dissemination of evidence for this target audience, including, for example: support to health policymakers for decision-making in health policies and programs, the organization of services and /or health systems, encouraging the use of scientific evidence in the decision-making process, the access to health information from the population perspective, the adaptation of knowledge obtained from evidence to the local context, among others.
- C (context): we will include any strategy related to health evidence communication, at the individual or population level, in the context of public or supplementary health, at any level of care (health unit, neighbourhood, municipality, state, region or country).

We will consider any primary (descriptive or analytical) or secondary study design addressing strategies for communicating health evidence will be considered, both for managers and for the population, in the context of public or private health.

### Search strategy

We will conduct a comprehensive, sensitive, and unrestricted search in the literature through structured search strategies, with relevant descriptors and synonyms, for the following databases:

- Campbell Collaboration
- Cochrane Library (via Wiley)
- EMBASE (via Elsevier)
- Biblioteca Virtual em Saúde (BVS)
- Epistemonikos
- Health Evidence
- Health Systems Evidence
- International Bibliography of the Social Sciences (IBSS)
- Medical Literature Analysis and Retrieval System Online (MEDLINE, via PubMed)
- PDQ-Evidence (https://www.pdq-evidence.org/)
- Rx for Change (https://www.cadth.ca/rx-change)

Additional unstructured searches will be carried out on the following sources of evidence-informed policies:

- Agency for Healthcare Research and Quality AHRQ/EUA (www.guidelines.gov)
- Centre for Reviews and Dissemination, Service Delivery and Organization (https://www.york.ac.uk/crd/research/service-delivery/)
- Cochrane Effective Practice and Organization of Care (EPOC) (https://epoc.cochrane.org/)
- EPPI-Centre (https://eppi.ioe.ac.uk/cms/Default.aspxãtabid=56)
- European Observatory on Health Systems and Policies (https://eurohealthobservatory.who.int/)
- Evidence Informed Policy Networks (EVIPnet)
- International Initiative for Impact Evaluation (3ie) (https://www.3ieimpact.org/)
- McMaster University’s Health Forum (https://www.mcmasterforum.org/)
- Supporting the use of Research Evidence (SURE) (https://epoc.cochrane.org/sites/epoc.cochrane.org/files/public/uploads/SURE-Guides-v2.1/Collectedfiles/sure_guides.html)
- The Alliance for Health Policy and Systems Research (https://ahpsr.who.int/)
- What Works Centres (https://www.gov.uk/guidance/what-works-network)

Electronic search will be performed in the following grey literature databases:

- Opengrey (https://opengrey.eu)
- ProQuest Dissertation and Theses (https://pqdtopen.proquest.com/)
- Thesis Commons (https://thesiscommons.org/)
- Open Access Theses and Dissertations (https://oatd.org/)

Electronic search will be performed in the following preprint repositories:

- Europe PMC (https://europepmc.org/)
- Open Science Framework (https://osf.io/preprints/)

Manual searches will be carried out in lists of relevant studies and contacts with content experts. No language filter will be applied. The search will be restricted to the period from 2000, considering the advances and changes in digital and social media that have taken place mainly in the last two decades. Full publications or abstracts presented at congresses and events will be included.

### Study selection process

The study selection process will be carried out in two phases using the Rayyan platform^6^. The first phase will consist of the titles and abstracts screening all the references retrieved by the search strategies. Then, references will be categorized as “potentially eligible” or “eliminated”. The second phase will consist of analyzing the full text of the “potentially eligible” studies to confirm their eligibility or exclude them in the second phase (justifications for each exclusion in the second phase will be presented). Two groups of independent reviewers will carry out the two selection phases, and a third reviewer will solve disagreements in decisions to include or exclude studies. The entire selection process will be presented through a PRISMA flowchart.

### Data extraction

Two reviewers will independently extract data from communication strategies or programs identified in the included studies, and differences in information register will be solved by consulting a third reviewer. The following data will be collected for each included study:

- Proposing institution (governmental, non-governmental, others).
- Country/locality for which the strategy was developed.
- Target audience: health policymakers, population, both.
- Strategy or program focus: individual, group or community.
- Proposed scenario for execution: public or private health.
- Strategy content: single or multiple interventions.
- Duration of the strategy (perennial or temporary).
- Strategy status: proposed, executed, and not evaluated, executed, and evaluated.
- Expected costs for implementing the strategy.
- The impact expected by proponents (outcomes).
- Barriers and facilitators identified for strategy implementation.

Authors of included studies may be contacted if additional information is required.

### Methodological quality assessment (risk of bias) assessment of the included studies

The purpose of this scoping review is to map the strategies to communicate health evidence presented in descriptive studies or use cut-offs of analytical studies that report strategies related to health evidence communication. Thus, the included studies methodological quality assessment will not be applied, as recommended by the Joanna Briggs Institute for scoping reviews^4^.

### Data synthesis and presentation of the results

The strategies related to health evidence communication will be classified using the categories defined based on the data described above. This qualitative synthesis will be presented using a narrative approach and in graphics and/or tables. Depending on the availability of information, descriptive statistics will be performed using Microsoft Excel® and/or STATA® software.

In the end, we will propose a product to communicate the results of this scoping review to users of the obtained information.

## Data Availability

All data produced in the present study are available upon reasonable request to the authors

## Acknowledgements

We would like to acknowledge the contribution of the ESPIE (Apoio à formulação e implementação de políticas de saúde informadas por evidências) project team: Silvio Fernandes da Silva, Davi Mamblona Marques Romão, Jorge Otávio Maia Barreto, Maria Lúcia Teixeira Machado and Romeu Gomes in the early stage of this protocol.

## References

Brownell SE, Price JV Steinman L et al (2013) Science communication to the general public: why we need to teach undergraduate and graduate students this skill as part of their formal scientific training. J Undergrad Neurosci Educ 12:E6–E10.

Fontaine G, Lavallée A Maheu-Cadotte MA (2018) Health science communication strategies used by researchers with the public in the digital and social media ecosystem: a systematic scoping review protocol. BMJ Open 30;8(1):e019833.

McCormack L, Sheridan S Lewis M et al (2013) Communication and dissemination strategies to facilitate the use of health-related evidence. In: Database of Abstracts of Reviews of Effects (DARE): Quality-assessed Reviews [Internet]. York (UK): Centre for Reviews and Dissemination (UK); 1995-. Available from: https://www.ncbi.nlm.nih.gov/books/NBK174174/.

Ouzzani M, Hammady H Fedorowicz Z et al (2016) Rayyan-a web and mobile app for systematic reviews. Syst Rev 5:1–11.

Peters MDJ, Godfrey C McInerney P et al (2020) Chapter 11: Scoping Reviews (2020 version). In: Aromataris E, Munn Z (eds) JBI Manual for Evidence Synthesis. JBI.

Tricco AC, Lillie E Zarin W et al (2018) PRISMA extension for scoping reviews (PRISMA-ScR): checklist and explanation. Ann Intern Med 169(7):467–473.

